# Functional profiling of human chorionic gonadotrophin in embryo peri- and post-implantation *in vitro* models

**DOI:** 10.64898/2026.04.01.26349947

**Authors:** Darja Lavogina, Apostol Apostolov, Sanjiv Risal, Paula Iglesias Moreno, Amruta D. S. Pathare, Anette Roop, Mathilde Bergamelli, Ilmatar Rooda, Kristina Hansing, Merli Saare, Fredrik Lanner, Ganesh Acharya, Jennifer J. Adibi, Pauliina Damdimopoulou, Alberto Sola-Leyva, Hannu Koistinen, Andres Salumets

## Abstract

Human embryo implantation, occurring approximately one week after fertilization, remains poorly understood due to ethical and technical limitations of *in vivo* investigation. To overcome these barriers, and model this critical developmental event, encompassing peri- and early post-implantation stages, we used an *in vitro* embryo attachment model composed of donor-derived endometrial epithelial cells forming an open-faced endometrial layer (OFEL) and human stem cell-derived blastoids recapitulating human day 5 blastocysts in peri-implantation model. Following attachment, developmental progression was further investigated on laminin-coated substrates to capture early post-implantation dynamics. Despite its central role as the primary endocrine signal of early pregnancy, human chorionic gonadotropin (hCG) remains largely uncharacterized in this context. Here, we describe the transcriptomic profile of blastoid–endometrial co-cultures relative to OFEL alone, identifying *CGA* and *CGB3/5/8* as among the most strongly upregulated genes following blastoid attachment to hormonally stimulated OFEL. Consistent with these findings, immunoassays and luteinizing hormone/choriogonadotropin receptor (LHCGR) activation assays of conditioned media confirmed the secretion of heterodimeric, biologically active hCG and its free subunits in co-cultures, but not in endometrial layers alone. Notably, the hyperglycosylated hCG heterodimer was the predominant isoform detected. Co-culture with the endometrial component significantly increased hCG secretion compared with blastoids cultured alone, an effect further enhanced by hormonal priming in the peri-implantation model. Collectively, these findings indicate that a hormonally primed endometrial environment not only promotes blastoid attachment but also amplifies embryonic hCG production and bioactivity, underscoring the importance of maternal endocrine cues in early embryo-endometrium communication. Furthermore, our peri-and early post-implantation models recapitulate key aspects of reciprocal endocrine signaling between embryonic and endometrial tissues, providing a tractable experimental framework to investigate embryo–endometrium crosstalk.

## 1. Introduction

The process of human embryo implantation represents a critical developmental milestone that establishes pregnancy. However, its investigation in humans remains severely constrained by major ethical and technical challenges. Limited access to early human embryos, together with the inaccessibility of the implantation site deep within the maternal endometrium, has long hindered direct analysis of the cellular and molecular mechanisms underlying this event. Consequently, much of our current understanding derives from animal models, which differ substantially from humans in endometrial physiology, embryo implantation dynamics, and placentation [1,2].

Recent advances in stem cell and organoid technologies have enabled the reconstruction of key aspects of human implantation *in vitro*, providing ethically acceptable and experimentally tractable systems [3,4]. Naïve human embryonic stem cells (hESCs) can self-organize into blastocyst-like structures, termed blastoids [5], which recapitulate early embryonic architecture and lineage segregation into the epiblast (EPI), hypoblast (primitive endoderm, PrE), and trophectoderm (TE) according to the timing of blastocyst development [6,7]. In parallel, endometrial epithelial organoids (EEOs), derived from primary tissue, reproduce key physiological and morphological features of the uterine epithelium, including responsiveness to hormonal cues [8]. When cultured as two-dimensional open-faced endometrial layers (OFELs), epithelial cells expose their apical surface, enabling direct interaction and attachment of blastoids [3], thereby providing a physiologically relevant platform to model embryo-endometrium crosstalk during implantation.

Following attachment, early post-implantation development is characterized by rapid embryonic reorganization, trophoblast differentiation, and activation of endocrine signalling. *In vitro* studies demonstrate that attachment initiates coordinated morphogenetic and transcriptional programs in human embryos and blastoids that parallel in vivo lineage patterning and early gastrulation, establishing attachment as a developmental trigger rather than a purely physical event [9,10]. To capture post-implantation progression, laminin-521-coated substrates have been used to specifically mimic the human endometrial basement membrane [11,12]. This environment supports integrin-mediated interaction (particularly via α6β1 and α3β1 expressed in EPI and TE) essential for development following initial implantation [12]. While OFEL-based co-culture models recapitulate peri-implantation events, simplified laminin-coated systems enable longitudinal analysis of trophoblast differentiation and endocrine function, such as human chorionic gonadotropin (hCG) synthesis, over extended 13-day culture period.

Among the earliest molecular signals exchanged between the implanting embryo and the maternal endometrium is hCG, the principal endocrine hormone of pregnancy [13]. hCG is a heterodimer composed of an α-subunit (hCGα, encoded by *CGA*), shared with other glycoprotein hormones [14], and a β-subunit (hCGβ) encoded by a cluster of closely related nonallelic genes (*CGB1*, *CGB2*, *CGB3*, *CGB5*, *CGB7*, and *CGB8*). In blood or urine of a pregnant female, hCG exists as a biologically active intact heterodimer and as a hyperglycosylated variant (hCG-h), with slightly lower affinity to the luteinizing hormone/chorionic gonadotropin receptor (LHCGR) [15]. Free hCGα and hCGβ are also detectable in urine. The relative abundance of these forms changes dynamically across early pregnancy [16], with hCG-h dominating during implantation and early placentation, followed by the rise of “regular” hCG during luteal support and later gestation. Classically, hCG exerts its effects through activation of the LHCGR, stimulating cAMP-dependent progesterone production in the corpus luteum, which is essential for pregnancy maintenance [17]. However, emerging evidence suggests that hCG heterodimer and hCGβ may also exert LHCGR-independent actions [18,19], including the regulation of trophoblast differentiation, indicating a broader role for this hormone in the maternal-embryonic interface.

Recent studies have shown that human embryos and blastoids can attach to and interact with endometrial cells in *vitro*, enabling molecular interrogation of implantation and early post-implantation events, including trophoblast invasion and lineage specification [20,21]. Although embryonic hCG secretion has been implicated in early embryo-endometrium communication in such models [22], its regulation, bioactivity, and modulation by the endometrial environment are not yet fully elucidated.

Here, we established and characterized an *in vitro* implantation platform combining donor-derived endometrial epithelial cells with hPSC-derived blastoids to investigate hCG-mediated embryo-endometrium interactions. The experimental workflow (Figure 1) integrates blastoids generation, hormonal priming of OFELs to mimic receptive endometrium, and co-culture to enable blastoid attachment. In parallel, a complementary two⍰dimensional laminin-based system was used to model early post-implantation development and to monitor trophoblast differentiation and hCG production over a 1311day period. These models were analysed by immunofluorescence and transcriptomics to capture cellular responses, alongside immunoassays and functional bioassays of conditioned media to quantify hormone secretion and biological activity. By focusing on hCG as a readout of embryonic endocrine function, and glycodelin as a marker of endometrial receptivity, this study provides a robust and tractable framework to dissect the molecular dialogue underlying human peri- and early post-implantation development.

**Figure 1.**
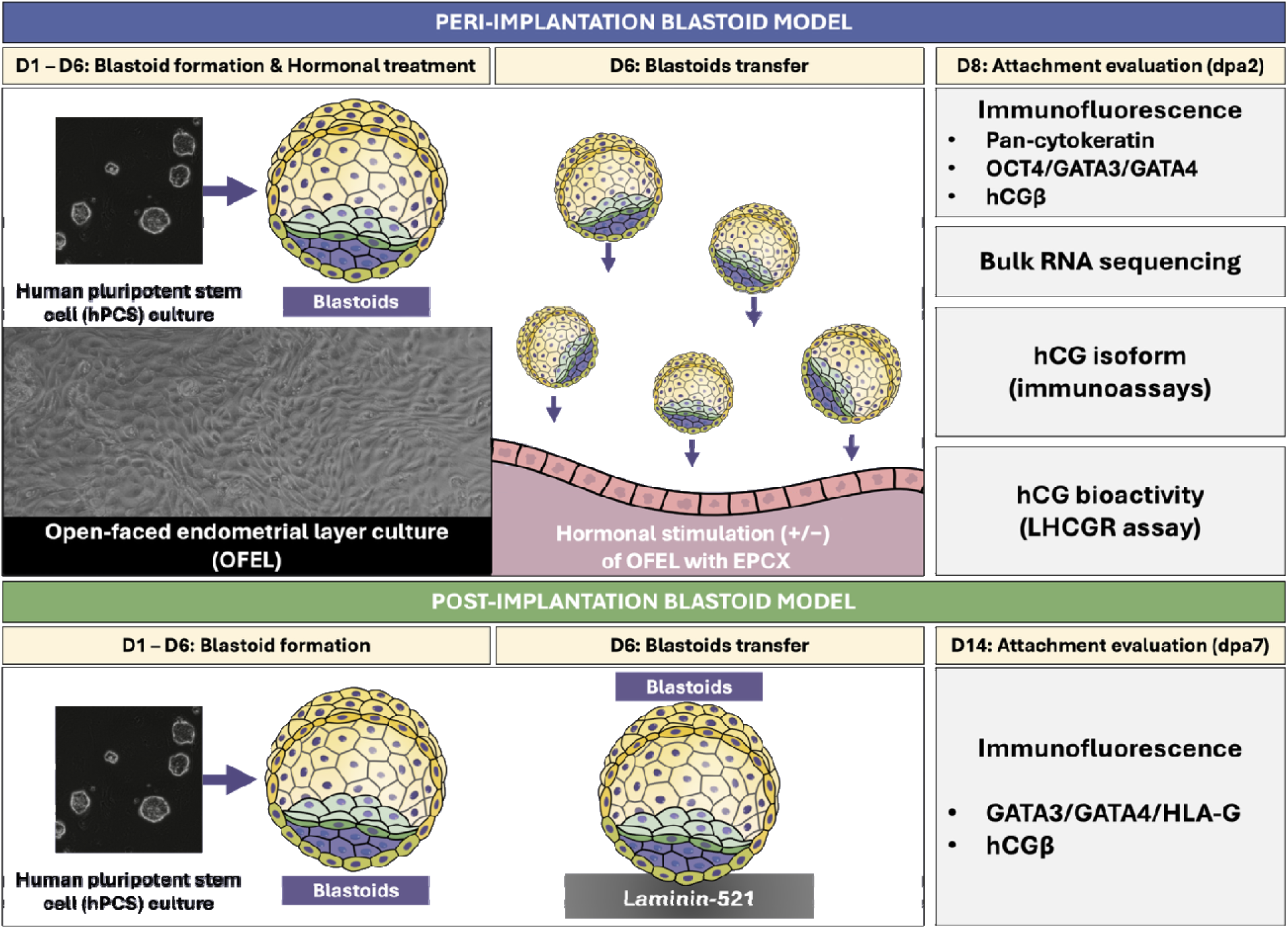
Experimental workflow; days in culture are shown in the top part of the chart. OFEL, open-faced endometrial layer; EPCX, 10 nM oestrogen (E2), 1 μM progesterone (P4), 250 μM 8-Br-cAMP, and 10 μM XAV-939; dpa, day post-attachment; hCG, human chorionic gonadotropin; LHCGR, luteinizing hormone/choriogonadotropin receptor; hPSCs, human pluripotent stem cells.

## 2. Materials and methods

### 2.1. Blastoid culture

Blastoid generation was performed as previously described. Briefly, naïve H9 hESCs (46,XX cell line from (WiCell Research Institute, Madison, Wisconsin) were cultured on feeder layer, mouse embryonic fibroblast (MEF), in PXGL medium (N2B27 supplemented with the mitogen-activated protein kinase kinase inhibitor PD0325901, tankyrase inhibitor XAV-939, protein kinase C inhibitor Gö 6983 and hLIF) under hypoxic condition (5% CO□, 5% O□, 90% N_2_) [23]. For blastoid generation, naïve H9 cells were dissociated with Accutase (Gibco), pelleted, and resuspended in PXGL plus Rho kinase inhibitor Y-27632 (STEMCELL Technologies, Canada), then pre-plated for 1 h to remove MEFs. After counting, cells were transferred to aggregation medium and seeded at 100,000 cells per well in AggreWell 400 plates (STEMCELL Technologies, Canada). Aggregates formed for 24 h (day 1). On day 2, 2× PALLY medium was added without prior medium removal, resulting in a final concentration of 1× PALLY due to dilution with the existing AggreWell medium. On day 3, the medium was refreshed by removing the existing medium and replacing it with a full volume of fresh 1× PALLY [6]. On day 4, cultures were switched to N2B27 containing lysophosphatidic acid, bovine serum albumin and Y-27632. Blastoids were assessed on day 5 and collected on day 6.

### 2.2. Collection of endometrial tissue samples

Endometrial biopsies were obtained from healthy, fertile women (donor 1: age 34, BMI 28; donor 2: age 28, BMI 20.1) using a Pipelle flexible suction catheter (Laboratoire CCD, France) during the early secretory phase (day 3 after the luteinizing hormone surge). Fertility was confirmed by a history of at least two live births. The donors had not used hormonal medications for at least three months prior to biopsy collection and did not have any history of sexually transmitted infections, uterine disorders, endometriosis, or polycystic ovary syndrome. Endometrial tissues were transported in HypoThermosol FRS preservation solution (Sigma-Aldrich, USA).

### 2.3. Endometrial epithelial organoid culture

EEOs were generated as previously described [8]. Endometrial tissue samples were dissociated enzymatically with 1.25 U/mL dispase II (Sigma-Aldrich, USA) and 0.4 mg/mL collagenase V (Sigma-Aldrich, USA) at 37 °C for up to 20 min. The digested suspension was filtered through a 100-μm cell strainer, which was subsequently backwashed to collect the gland-enriched fraction of epithelial cells. This fraction was then seeded in 48-well plates (Greiner Bio-One, Austria) by resuspending in a 25 µL drop of Matrigel (Corning, USA) and cultured at 37 °C in 250 µL expansion medium for up to 10 days before passaging or freezing according to the previously reported protocols [8,24]. The media were replaced every 2-3 days, and the organoids were passaged every 7-10 days.

### 2.4. Open-faced endometrial layer generation and hormonal stimulation

To promote attachment of the primary epithelial cells, 96-well plates (Revvity, USA) were pre-coated with 3% Matrigel (Corning, USA) diluted in the Advanced DMEM/F12 medium (Gibco, USA) and allowed to solidify for 2-3 h. The EEOs were dissociated into single cells and seeded in a 2D monolayer (OFEL) for the attachment assay. To remove Matrigel, EEOs were incubated with 25 μL of dispase II (10 mg/mL) (Sigma-Aldrich, USA) at 37 °C for 1 h. Further, the EEOs were transferred to a 1.5 mL low-binding tube and centrifuged at 600 x *g* for 5 minutes. After washing with Advanced DMEM/F12, the EEOs were resuspended in the dissociation solution containing 1 mL of TrypLE (Fisher, UK), 51 μg/mL N-acetylcysteine (Merck, Germany) and 10 μM Y-27632 (STEMCELL Technologies, Canada), and incubated at 37 °C for 15 min, resuspending the mixture every 5 minutes until the EEOs were dissociated to single cells. After centrifugation, the epithelial cells were counted and seeded at a density of 25,000 cells per well in a 96-well plate (Revvity, USA) or 100,000 cells per well of an 8-well chamber (Corning, USA). The hormonal stimulation of OFEL was performed using 10 nM oestrogen (E2; Sigma-Aldrich, USA), 1 μM progesterone (P4; Sigma-Aldrich, USA), 250 μM 8-Br-cAMP (Tocris, UK) and 10 μM XAV-939 (STEMCELL Technologies, Canada) for 6 days. Conditioned culture media were collected and stored at -80 °C for subsequent hCG analyses (see section 2.8).

### 2.5. Peri- and early post-implantation models

The attachment assay was done by transferring five day-6 blastoids per OFEL-containing well with a stripper pipette (CooperSurgical, USA). At this stage, the medium was exchanged to attachment medium (CMRL supplemented with hormones), containing embryonic stem cell–qualified FBS (10%; Gibco), GlutaMAX (2 mM; Gibco), N2 supplement (1×; Gibco), B27 supplement (1×; Gibco), sodium pyruvate (1 mM; Gibco), E2 (10 nM; Sigma-Aldrich), P4 (1 µM; Sigma-Aldrich), and Y-27632 (10 µM; STEMCELL Technologies). A hormone-free CMRL control medium containing the same supplements, but lacking E2 and P4 was used in parallel.

After 2 days, the percentage of attached blastoids relative to the total number of blastoids transferred was assessed by washing each well twice with CMRL medium and counting the number of attached blastoids per well [3]. Subsequently, conditioned culture media were collected and stored at -80 °C until further hCG detection (see sections 2.8 and 2.9), and the cellular fraction of the model was either lysed for RNA extraction and bulk RNA sequencing (see section 2.7) or fixed for immunostaining (see section 2.6). For the conditioned media samples collected in the absence of the OFEL, five blastoids were transferred to the ultra-low attachment plate in the same medium and grown for the same time as the blastoids in the presence of OFEL.

For post-implantation analyses, individual blastoids (one blastoid per well) were selected based on morphology and placed into 8-well chambered slides (Corning, USA) coated with laminin-521 (#LN521-02; BioLamina, USA) at a concentration of 10 µg/ml[10,22]. The blastoids were first grown according to the previously published protocols [9,25] in IVC1 medium composed of 50% DMEM/F12 and 50% Neurobasal medium supplemented with 20% heat-inactivated fetal bovine serum (Invitrogen, USA), 1× GlutaMAX (Gibco, USA), 1× ITS-X (Gibco, USA), 8 nM E2 (E2; Sigma-Aldrich, USA), 200 ng/mL P4 (P4; Sigma-Aldrich, USA), and 25 mM N-acetyl-L-cysteine (Merck, Germany). At day 2 post-attachment (dpa2), the medium was changed to IVC2 and physioxic conditions (5% CO□, 5% O□, 90% N_2_) were applied. This medium also contained 50% DMEM/F12 and 50% Neurobasal, plus 30% KnockOut Serum Replacement, 1× GlutaMAX, 8 nM E2, 200 ng/mL P4, and 10 μM Y-27632. The IVC2 medium was replaced every other day. The post-implantation culture was maintained until 7 days post-attachment (dpa7).

### 2.6. Immunofluorescence experiments

The OFEL, blastoids and peri- and post-attachment models were fixed in 4% paraformaldehyde (PFA) in phosphate-buffered saline (PBS) (Gibco, USA) and permeabilized by washing three times with 0.01% Tween-20 (Sigma-Aldrich, USA) in PBS without calcium and magnesium (Qiagen, Germany). Samples were then blocked with bovine serum albumin (BSA, 5% w/v, Sigma, USA) and normal donkey serum (20% v/v) in PBS (Sigma-Aldrich, USA) was applied for 30 minutes at 4 °C. The incubation with the primary antibodies took place overnight at 4 °C; the antibodies were diluted in washing buffer containing 0.1% Triton X-100 (Sigma-Aldrich) and 0.2% BSA in PBS. The OFEL was immunostained with mouse anti-human pan-cytokeratin antibody (ab86734; Abcam, UK) at 1:50 dilution. Blastoids were stained with mouse anti-human Octamer Transcription Factor 4 (OCT4) antibody for EPI (sc-5279; Santa Cruz, Germany), rabbit anti-human GATA-binding protein 3 (GATA3) antibody for TE (sc-9009; Santa Cruz, Germany), rat anti-human GATA-binding protein 4 (GATA4) antibody for PrE (14-9980-82; eBioscience™, USA), mouse monoclonal antibody against hCGβ as a free subunit or in the context of heterodimer (ab9582; Abcam, UK), each at 1:100 dilution; alternatively, human leukocyte antigen-G (HLA-G) antibody (4H84; sc21799, Santa Cruz, USA) at 1:200 dilution. After washes, incubation with the secondary antibodies (all at 1:1000 dilution) was carried out at room temperature for 1 h. Goat anti-mouse IgG antibody labelled with Alexa 488 (A-11001) or donkey anti-mouse IgG (H+L) labelled with Alexa Fluor™ 555 (A-31570), goat anti-rabbit IgG (H+L) labelled with Alexa Fluor™ 546 (A-11035) and donkey anti-rat IgG (H+L) labelled with Alexa Fluor™ 647 (A48272TR; all from Invitrogen, USA) were used. 4′,6-diamidino-2-phenylindole (DAPI, Invitrogen, USA) was used at 1:1000 dilution in the washing buffer to stain the nuclei of the cells. Images were taken with 10×, 20× or 25× objectives using a multipoint spinning disk confocal Nikon Ti2 microscope (Nikon, Japan).

### 2.7. RNA extraction, library preparation and sequencing

RNA was extracted using the miRNeasy Tissue/Cells Advanced Micro Kit (Qiagen, Germany), and RNA quality and quantity were assessed using the Agilent 2100 Bioanalyzer with RNA ScreenTape (Agilent, USA). The isolated RNA was stored at -80 °C until further analyses. The library was prepared using 200 ng input RNA with TruSeq Stranded mRNA kit (Illumina, USA) and sequenced on NextSeq 1000 (Illumina, USA) using single-end 80-bp reads.

### 2.8. Immunoassays for glycodelin and different forms of hCG and its subunits

Prior to the measurement of the different forms of hCG, conditioned media samples were diluted to the volume ratio of 1:30 to 1:50 with DELFIA Buffer (Revvity/Wallac, Finland). For glycodelin, the samples were diluted to the volume ratio of 1:3 in the assay buffer containing 50 mM Tris, pH 7.7, 0.9% NaCl, 0.05% NaN_3_, 0.5% BSA, 0.05% alfa-globulins and 10 mg/500 ml diethylenetriaminepentaacetic acid (DTPA). The total hCG heterodimer levels were detected using a commercial immunoassay (DELFIA hCG kit, Revvity/Wallac; analytical sensitivity of 0.5 IU/L) according to the manufacturer’s instructions. The other forms of hCG and glycodelin were detected using the in-house immunoassays according to the previously reported protocols: free hCGα with limit of detection (LoD) of 2.8 pmol/L [26], free hCGβ with LoD of 0.3 pmol/L [27,28], hCG-h with LoD of 2 pmol/L [29], and glycodelin with limit of quantitation < 1 ng/mL [30]. The assay for hyperglycosylated hCG detects, in addition to the hCG-h, also hyperglycosylated hCGβ, but the response is *ca* 25% of that for hCG-h [31].

### 2.9. LHCGR activation assay

For monitoring activation of LHCGR, Madin-Darby Canine Kidney (MDCK) cells stably expressing human LHCGR were used (a kind gift from Dr Prema Narayan, Southern Illinois University). The cells were grown as adherent monolayers at 37□°C and 5% CO_2_ in a humidified incubator (Sanyo, Japan) in Dulbecco’s Modified Eagle’s Medium and Ham’s F-12 nutrient mixture (Corning, USA) supplemented with 10% fetal bovine serum, 100 U/mL penicillin, 0.1 mg/mL streptomycin and 0.25 μg/mL amphotericin B (all from Capricorn Scientific GmbH, Germany). For measurement of intracellular cAMP, the cells were transduced with the biosensor expression vector mTurquoise2Δ-Epac(CD, ΔDEP, Q270E)-td^cp173^Venus (H187; [32]) cloned into the BacMam system-compatible vector as previously described [15,33].

The assay was carried out according to the previously reported protocols [15,33] with minor modifications. Briefly, cells were treated with viral stock (multiplicity of infection: 100-300) in 5□mL of growth medium for 3-5□h□at 37□°C in a humidified CO_2_ incubator. Afterwards, the medium was aspirated, the cells were trypsinized and resuspended in fresh growth medium containing 10 mM sodium butyrate (Sigma-Aldrich, USA). Next, the cells were seeded onto the cell culture-treated black 96-well clear-bottom plates (Corning #3340, USA) at the density of about 15,000-20,000□cells per well and cultured for further 26-44□h at 37□°C. The growth medium was replaced with 90□μL of PBS supplemented with Ca^2+^ and Mg^2+^ (Corning, USA) 20-30 min before the experiment.

Recombinant hCG (Ovitrelle) was used as a positive control (Merck Serono Europe, Germany). The 3-fold dilutions of Ovitrelle (final concentration ranging from 3 nM to 4.1 pM) or conditioned media (starting from 1:6 dilution) were made on transparent 96-well clear-bottom plates (269620, Thermo Fisher Scientific, Denmark) into Ca^2+^ and Mg^2+^-supplemented PBS containing 7.5□μmol/L BSA (Fisher Scientific, Belgium) to reduce non-specific binding; 7.5□μmol/L BSA in PBS was used as the negative control. To initiate LHCGR activation, 10 μL of the diluted Ovitrelle or conditioned media were added onto the cells. The fluorescence intensities were registered prior to and after addition of the diluted samples of interest using PHERAstar plate reader (BMG LABTECH GmbH, Germany), with excitation at 427□nm and simultaneous dual emission at 480 and 530□nm; readings were taken every 30 seconds for 60 minutes.

### 2.10. Data analysis and software

For general data analysis, GraphPad Prism version 10.6.0 (USA) and Excel version 2509 (Microsoft Office 365, USA) were used. For all methods, at least 2 independent experiments were carried out.

The bioinformatic analyses of transcriptomic data were carried out in the European Galaxy server [34]. The adapter and quality trimming was done using Trim Galore, followed by FastQC and MultiQC to ensure the quality of the files. Afterwards, HISAT2 was used to align the files to the reference genome (human genome GRCh38, version 43, Ensembl 109). The tool featureCounts aligned reads into the count table, and the differential expression analysis was performed with DESeq2. The differential gene expression for OFEL plus blastoid *versus* OFEL alone in the presence or absence of hormonal stimulation was filtered by P_adj_ value cut-off < 0.05 and binary logarithm of fold-change (log_2_FC) value cut-off ≤ -1 or ≥1. The short-listed differentially expressed genes (DEGs) are presented in the Supplementary Table S1. Venn diagram was generated using EVenn [35]. The analysis of gene ontology (GO) biological process enrichment was done with ShinyGO version 0.85 [36].

The human early embryogenesis reference dataset was obtained from the Early Embryogenesis Projection Tool [37] (version 2.1.2, available at https://petropoulos-lanner-labs.clintec.ki.se). UMAP coordinates and cell-type annotations were downloaded from the tool and used for visualization and for analysis of CGB cluster expression values.

The content of secreted hCG and glycodelin in conditioned media samples were calculated based on the immunoassay data from the corresponding calibration curves fitted using 4-parameter logistic model in WorkOut Plus software (Perkin-Elmer, USA). For samples where the signal was lower than 50% of LoD, the assigned hCG or glycodelin secretion value was 0. To facilitate the comparison of concentrations, total hCG concentrations expressed as IU/L were converted to pmol/L by multiplying by the factor of 2.93. To minimize the inter-experimental variation, the content of the same hCG form was normalized for each independent experiment and the OFEL donor according to the secretion obtained for the OFEL plus blastoids in the presence of hormones (average value set to 100%); in case of blastoids, the data was normalized analogously but averaged values obtained for both OFEL donors in the same experiment were taken into account. The normalized values were then pooled for the same hCG form and same type of conditioned media.

In LHCGR activation assay, the change in FRET ratio in time was calculated as reported previously [15,33], using fluorescence intensity values at 480 nm and 530 nm in each individual well before and after the addition of diluted samples of interest. To establish half-maximal effective concentration (EC_50_) for each sample of interest, ΔFRET values calculated for data points taken 60 min after the addition of diluted samples of interest were plotted against logarithm of concentration of the Ovitrelle control or against the logarithm of dilution factor of the sample (in case of conditioned media). To correct for high autofluorescence, in case of conditioned media samples additional correction was applied by subtracting ΔFRET value calculated for the blank OFEL medium sample from the ΔFRET value of the sample of interest. This correction was validated according to 2 independent experiments using blank OFEL medium alone, Ovitrelle in PBS, or Ovitrelle spiked into blank OFEL medium (see Supplementary Figure S1). The curves were analysed using the three-parameter logarithmic equation. In each independent experiment, the concentration of hCG in the samples of interest was calculated according to the EC_50_ of Ovitrelle and the EC_50_ of the conditioned media sample, assuming that the 50% activation of receptor is achieved at the same concentration of hCG in case of Ovitrelle and the conditioned media sample. The calculated values were then pooled for the same type of conditioned media.

## Results

### 3.1. Blastoids recapitulate embryonic lineage specification and enable peri- and post-implantation modelling

To evaluate lineage composition, generated blastoids were characterized by IF (Figure 2A). Prior to attachment, blastoids exhibited all three founding lineages, including GATA3-positive trophectoderm (TE), GATA4-positive hypoblast, and OCT4-positive epiblast (EPI) cells. To assess attachment capacity to the endometrial epithelial cells, blastoids were co-cultured with hormonally stimulated or unstimulated OFEL. In the peri-implantation model, bright-field imaging revealed that attached blastoids exhibited a flattened morphology, consistent with adhesion initiation, whereas non-attached blastoids retained a rounded morphology, indicative of the absence of adhesion (Figure 2B). Quantification of the attached blastoids demonstrated a significantly higher attachment rate in the hormonally stimulated OFEL compared to unstimulated conditions (Figure 2C).

**Figure 2.**
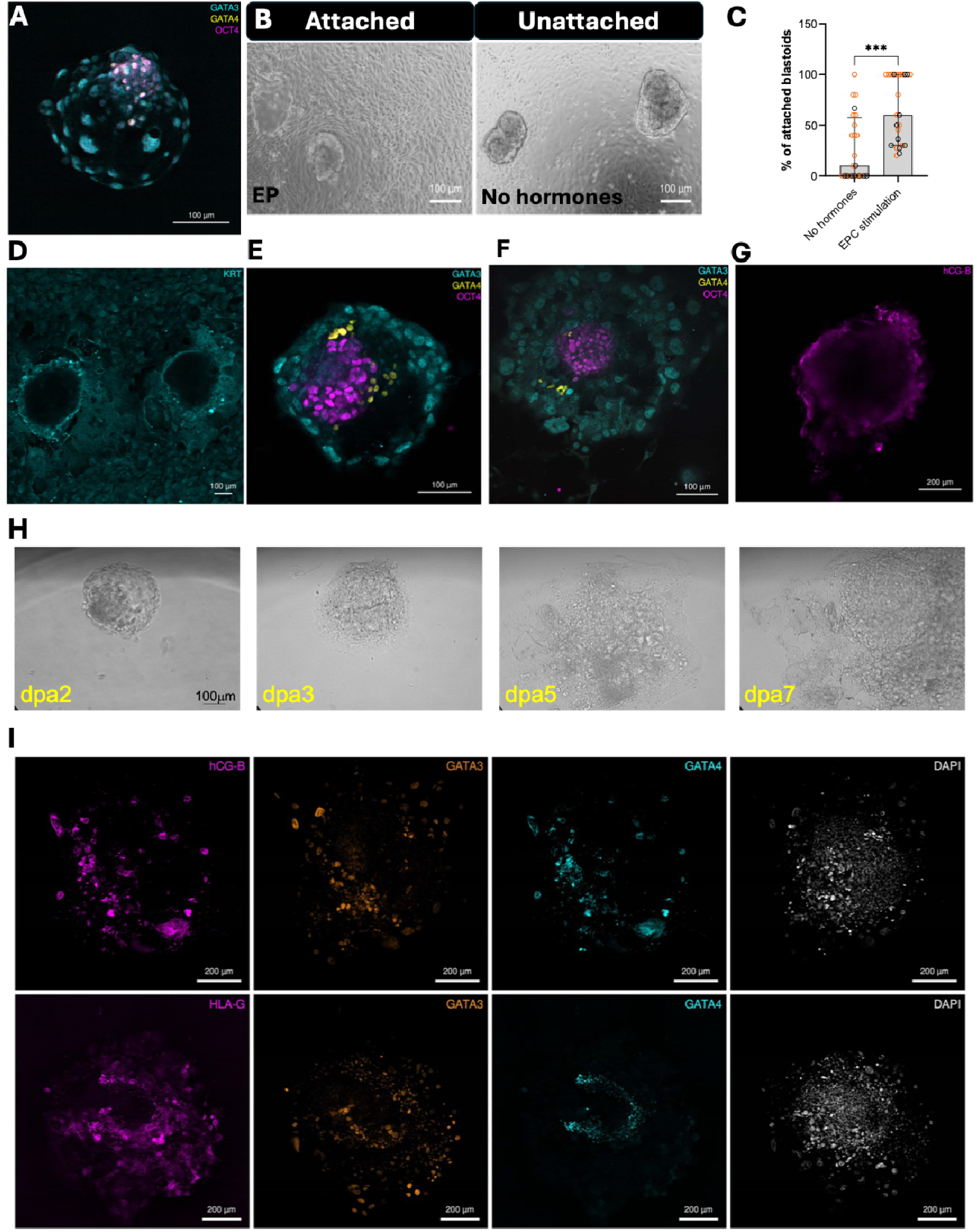
Peri- and post-implantation blastoid model. **A**, Characterization of the three cell lineages of human blastoids showing GATA3 (cyan; trophectoderm), GATA4 (yellow; hypoblast), and OCT4 (magenta; epiblast). **B**, Bright-field images of the model showing attached blastoid in hormonally stimulated (EP) open-faced endometrial layer (OFEL) conditions, and unattached blastoid in unstimulated OFEL conditions. **C**, The percentage of blastoids attached to OFEL, with and without hormonal stimulation, relative to the total amount of blastoids transferred is shown; column height features population median and error bars show quartiles. Each circle represents an independent experiment, and the colour of circles corresponds to two different OFEL donors. Statistical significance of pairwise comparisons (Mann-Whitney test): *** P < 0.001. **D**, Blastoids attached to the OFEL showing pan-cytokeratin staining (cyan; KRT). **E**–**F**, Blastoids 2 days post-attachment (dpa2) to hormonally stimulated (**E**) or unstimulated (**F**) OFEL showing GATA3, GATA4, and OCT4 staining. **G**, Blastoid attached to hormonally stimulated OFEL stained for hCGβ (magenta). Scale bars are provided at the bottom right corner of each image. **H**, Representative bright-field images of a post-attachment blastoid at dpa2 until dpa7. **I**, Immunostaining data showing cells positive for hCGβ (magenta), GATA3 (orange; trophectoderm), GATA4 (cyan; hypoblast), and DAPI (white; nucleus), and cells positive for HLA-G (magenta, extravillous trophoblast), in the post-attachment blastoids at dpa7. Scale bars are provided at the bottom right corner of each image

Following blastoid attachment, pan-cytokeratin staining was consistent with the epithelial identity of the OFEL and revealed localized adhesion at the blastoid–epithelial contact interface (Figure 2D). Furthermore, following attachment in the peri-implantation model, blastoids retained the three principal lineages on both hormonally stimulated and unstimulated OFELs (Figure 2E–F). To further assess trophoblast maturation, we stained blastoids attached to hormonally stimulated OFELs for hCGβ (Figure 2G). Expression of hCGβ, detected using a pan-hCGβ antibody recognizing both free and heterodimeric hCGβ, indicated trophoblast differentiation toward a syncytiotrophoblast (STB)-like fate.

To mimic post-implantation events in humans up to 7 days, we used a simplified *in vitro* model employing a laminin-521 coated two-dimensional culture system. Throughout this period, the post-attachment blastoids displayed progressive morphological transitions characteristic of post-implantation development, including structural reorganization and expansion from dpa2 to dpa7 (Figure 2H). Consistently, we observed the emergence of hCG-β positive STB as well as HLA-G positive extravillous trophoblast (EVT) cell populations by dpa7 (Figure 2I). By dpa7, post-attachment blastoids exhibited distinct trophoblast subtypes alongside organized lineage architecture, including GATA3-positive TE derivatives and GATA4-positive hypoblast cells (Figure 2I).

### 3.2. Transcriptomic profiling of the peri-implantation model reveals an in vivo-like early implantation signature

To elucidate the transcriptomic changes induced by the blastoid attachment in the peri-implantation model, we performed differential expression analysis comparing OFEL with blastoids *versus* OFEL alone in the presence or absence of the hormonal stimulation. The transcriptomics revealed 123 DEGs in the hormonally stimulated conditions comparison and 44 DEGs in the unstimulated conditions (Figure 3A, Supplementary Table S1). Among these, 41 genes overlapped between conditions, indicating a core transcriptional response to blastoid presence independent of hormonal stimulation. Notably, this shared gene set included *CGA* and *CGB3/5/8*, representing genes encoding the hCG α-subunit and the coding genes of the β-subunits, respectively (Figure 3B). Eighty-two DEGs were found exclusively in the hormonal treatment group, including *CGB7*, another gene encoding a β-subunit of hCG (Figure 3B). Consistently, normalized expression of the *CGB*-coding cluster (*CGB3/5/7/8*) and *CGA* demonstrated global upregulation of hCG-encoding genes upon hormonal stimulation (Figure 3C).

**Figure 3.**
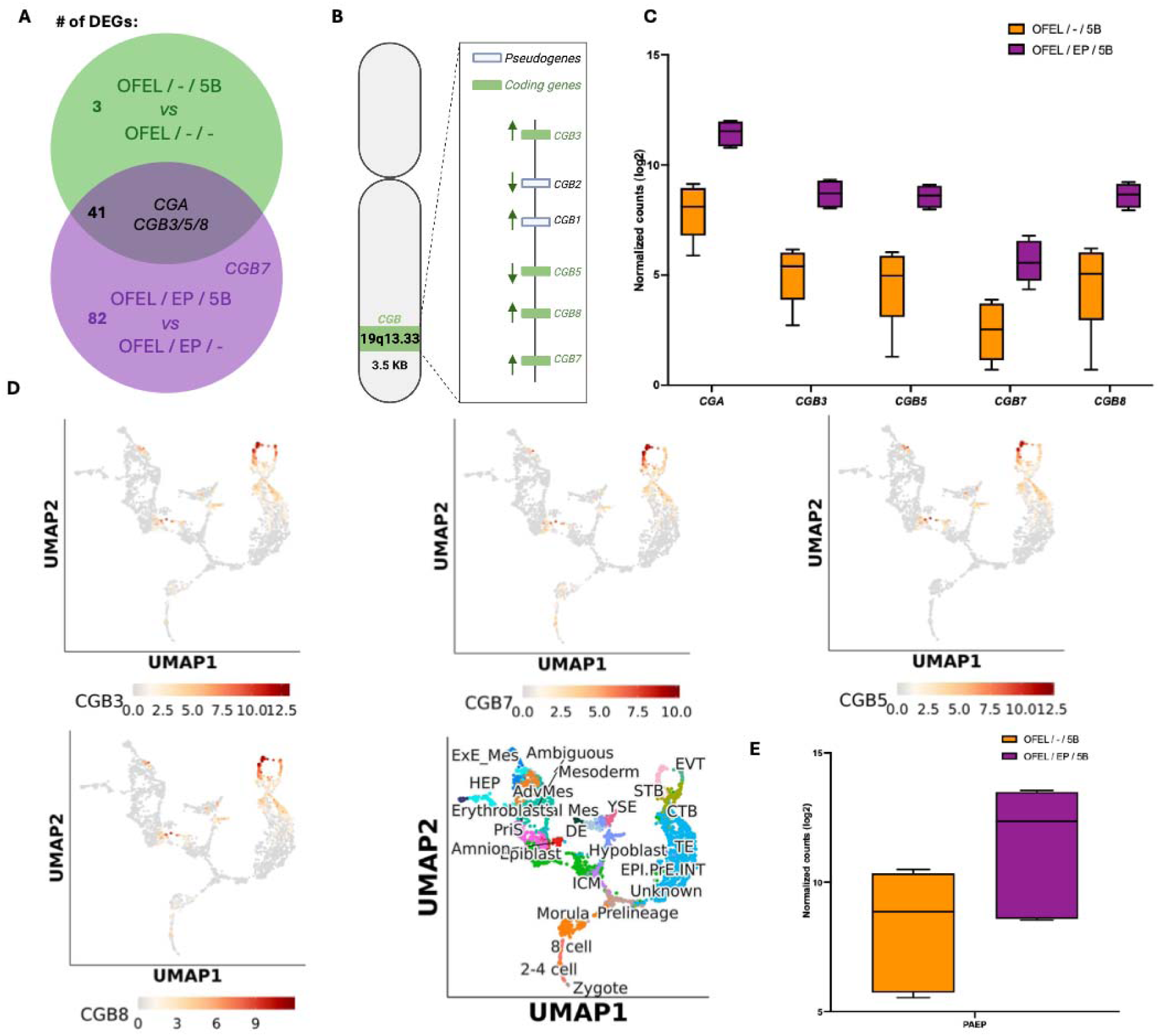
Differential expression analysis highlights peri-implantation transcriptional changes induced by the five blastoid (5B) attachment in hormonally stimulated (EP) or unstimulated (–) open-faced endometrial layer (OFEL). **A**, Venn diagram showing differentially expressed genes (DEGs) in the indicated comparisons; the total number of DEGs is shown on the left, and hCG-encoding genes identified as DEGs in the corresponding groups are indicated in italics. **B**, Diagram of the CGB gene cluster located at chromosome 19q13.33. Protein-coding genes are shown in green and pseudogenes in white. Arrowheads denote the direction of transcription for each gene. **C**, Normalized (log_2_-transformed) counts of coding genes in the CGB cluster and *CGA* in the attachment model (OFEL/–/5B (orange) and OFEL/EP/5B (purple)). **D**, UMAP projection of the single-cell embryo reference atlas with lineage annotations and expression of *CGB3*, *CGB5*, *CGB7*, and *CGB8*. **E**, Normalized (log_2_-transformed) counts of *PAEP* in the attachment model.

Furthermore, we assessed the expression of *PAEP* that encodes glycodelin – a glycoprotein secreted by the decidualized endometrium in the mid-secretory to late secretory phase of the menstrual cycle and an endometrial marker for receptivity [38,39]*. PAEP* expression was increased in the hormonally stimulated peri-implantation model, although this did not reach statistical significance (Figure 3E).

To better characterize the developmental dynamics of CGB expression, we examined CGB gene expression in a single-cell human embryo reference atlas spanning stages from the zygote to post-implantation embryos (Figure 3D). *CGB3/5/7/8* transcripts were detectable in the TE, cytotrophoblast (CTB), STB, and EVT, with highest expression observed in STB, consistent with its role as the primary source of hCG *in vivo*.

Accordingly, the gene ontology (GO) analysis of DEGs in the hormonally stimulated and unstimulated model highlighted biological processes associated with trophoblast differentiation and early implantation, including syncytium formation (Supplementary Figure S1) involving syncytin-1 protein that is encoded by *ERVW-1* and suppressing protein encoded by *ERVH48-1*) and maintenance of stem cell population (involving Sal-like protein 4 encoded by *SALL4*, T-box 3 encoded by *TBX3*, and PR domain zinc finger protein 14 encoded by *PRDM14* (Supplementary Table S1 A and B).

### 3.3. Immunoassays reveal secretion of multiple hCG isoforms by blastoids and modulation by the peri-implantation microenvironment

To assess the profile of hCG protein forms secreted by the blastoid in the peri-implantation model, in the presence or absence of OFEL or hormonal stimulation, immunoassays with conditioned culture media were carried out to detect the hCG heterodimer, its hyperglycosylated form, and free α- and β-subunits of hCG. Additionally, glycodelin was measured to evaluate endometrial responsiveness to hormonal stimulation.

For glycodelin (Fig. 4A), the highest levels were observed under hormonal stimulation with oestradiol, progesterone, cAMP and XAV-939 (EPCX). This was followed by the EP condition (oestradiol and progesterone) applied to OFEL-only cultures maintained for the same duration as in the attachment assay but in the absence of blastoids. These findings are consistent with the hormonal regulation of glycodelin and its role as a marker of endometrial epithelial receptivity. Glycodelin secretion appeared reduced in EP-stimulated OFEL in the presence of blastoids, although high inter-sample variability likely masked potential effects of co-culture. No glycodelin was detected in blastoid-only conditioned medium.

**Figure 4.**
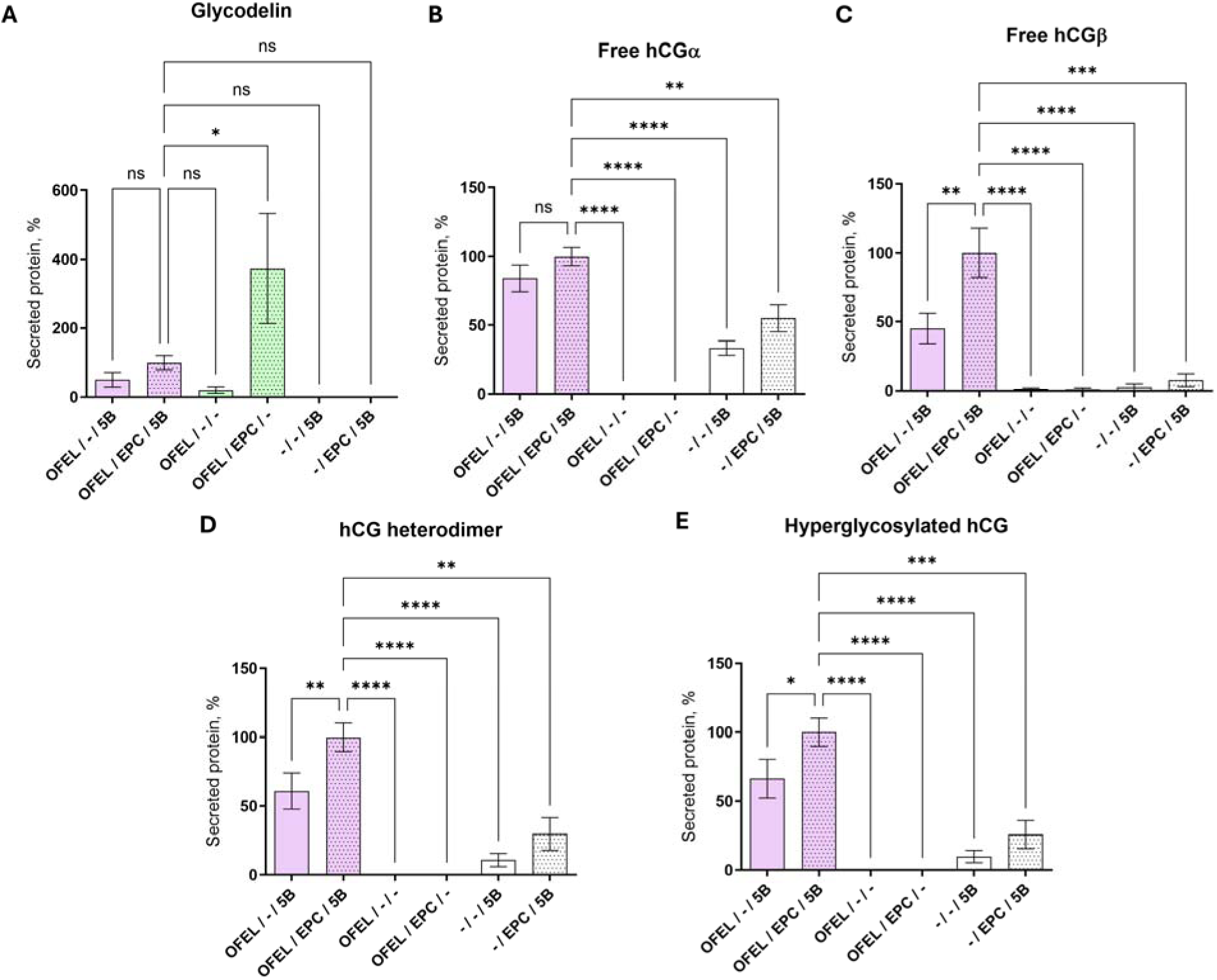
Secretion of protein markers in the conditioned culture media from peri-implantation model. Panels **A**-**E** feature immunoassay results in the peri-implantation model, the proteins of interest are listed above each graph and the sample treatment types under each graph (OFEL stands for open-faced endometrial layer, EP for hormonal stimulation and 5B stands for 5 blastoids). Average normalized secretion (levels for media representing the EP-stimulated OFEL and blastoid co-culture set to 100%) ± standard error of mean is shown for 2 independent experiments, each experiment using OFEL from 2 different donors. Statistical significance of grouped comparisons (ANOVA with Dunnett’s correction for multiple comparisons): **** P < 0.0001, *** P < 0.001, ** P < 0.01, * P < 0.05, ns – not significant.

In contrast, the profiles of hCG isoforms displayed the expected embryonic origin (Figure 4B–E). OFELs alone, whether unstimulated or EP-stimulated, showed no detectable secretion of hCG or its subunits. In co-culture condition of the peri-implantation model, attachment to hormonally stimulated OFEL significantly increased hCG secretion compared to blastoid-only cultures (P < 0.01), indicating enhanced embryo-endometrial crosstalk. All hCG forms, except hCGα, were further increased in the hormonally stimulated attachment model compared to non-hormonally stimulated conditions (P < 0.05), implying that a hormonally stimulated endometrial environment amplifies blastoid endocrine activity. The secretion profiles of total hCG and hCG-h were closely correlated (Supplementary Table S2), indicating that hyperglycosylated hCG represents the predominant isoform during peri-implantation stages.

Together, these findings showed that the hCG isoforms detected during early post-attachment stages blastoid are produced predominantly by trophoblast derivatives, particularly STBs and EVTs.

### 3.4. hCG secreted by attached blastoids is biologically active

To determine whether the hCG detected in the culture supernatants from peri-implantation model was biologically active, we assessed LHCGR-mediated signalling using a cell-based cAMP accumulation assay. Because the culture media exhibited high autofluorescence, we corrected all values by subtracting the background signal measured for the unconditioned media used for OFEL culture (considered as hCG-negative control). Due to lower throughput of this assay, media samples collected from the OFEL-only system were not analysed as these did not feature consistently detectable levels of hCG according to the immunoassay data.

Importantly, the presence of 8-Br-cAMP in the OFEL growth medium used prior to the attachment step did not interfere with the assay’s readout (Supplementary Figure S2). This conclusion was supported by the unchanged cAMP levels in spiked Ovitrelle controls and in media from blastoids cultured without OFEL. Given the substantial dilution of 8-Br-cAMP in the assay setup and its likely cellular uptake or partial hydrolysis during the co-culture period and the following storage of samples, any residual compound in the conditioned media was unlikely to contribute significantly to intracellular signalling in the reporter cells.

The results illustrating hCG/LHCGR axis activity are summarized in Figure 5. In most blastoid-only samples irrespective of the EP stimulation, LHCGR activation remained below the detection threshold, indicating minimal secretion of functionally active hCG. In contrast, blastoid-OFEL co-cultures consistently induced robust increase in intracellular cAMP, demonstrating the presence of biologically active hCG. Hormonal stimulation of OFEL (EP treatment) further increased active hCG secretion by the co-cultures, although this trend did not reach statistical significance according to ANOVA test; according to the paired Wilcoxon test, statistically significant difference (P < 0.05) was observed. Overall, the functional assay results mirrored the immunoassay data, confirming that the hCG produced within the attachment model was not only immunoreactive but also functionally able to activate LHCGR signalling.

**Figure 5.**
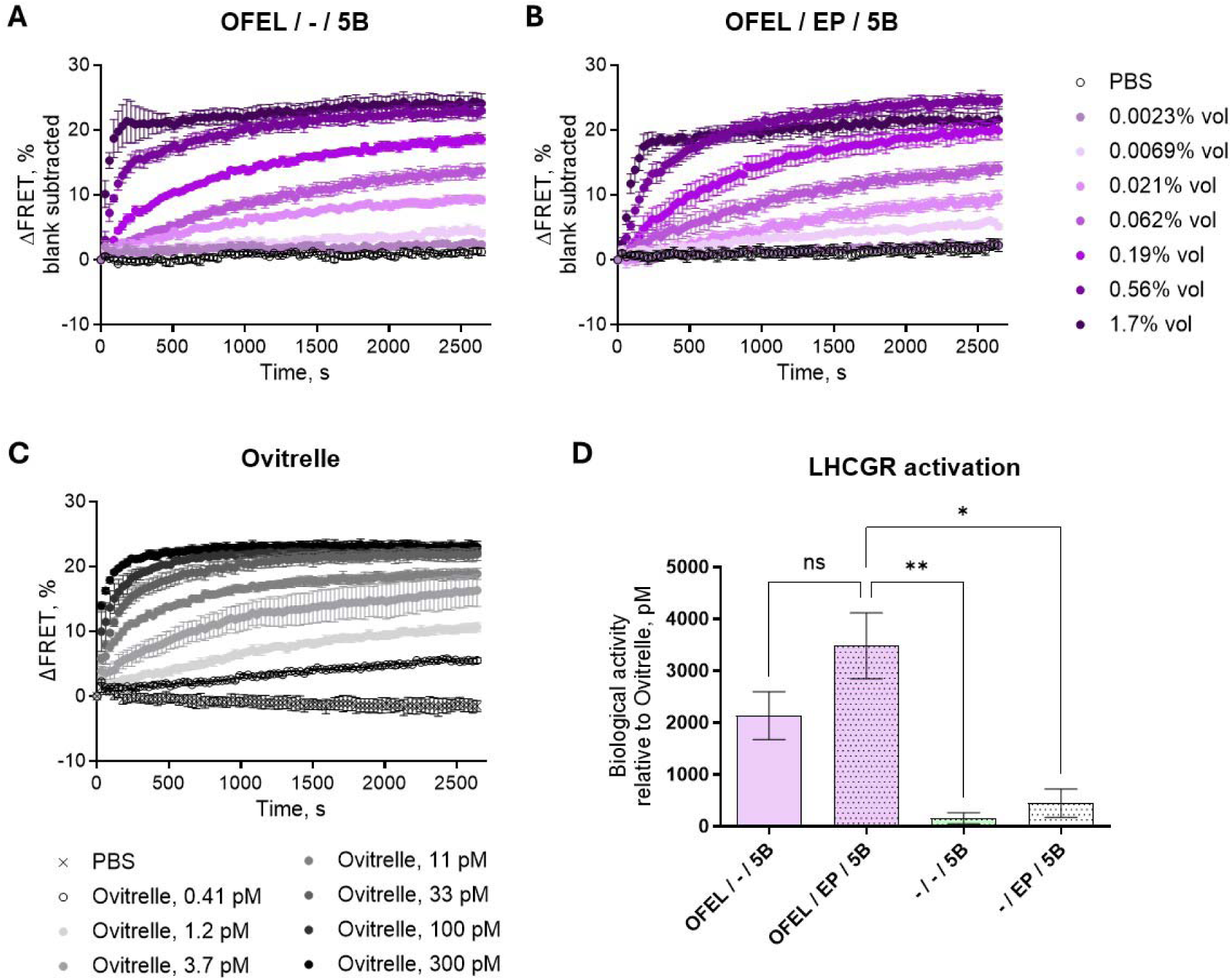
LHCGR activation assay in the peri-implantation model. **A-C**, Representative examples of cAMP-sensor signal change (ΔFRET) after addition of the hCG-containing samples at time t = 0 s. The type of treatment is indicated above the graph; for the conditioned medium samples (**A**–**B**), the dilutions used are indicated on the right side, and for the Ovitrelle control (**C**), the concentration of the recombinant hCG is listed under the graph. The kinetic curves show data from a single experiment; error bars indicate standard deviation of technical triplicates. **D**, Biological activity of hCG in conditioned culture media from the peri-implantation model. Based on the LHCGR-mediated increase in intracellular cAMP levels, hCG concentration in each sample was recalculated from comparison with recombinant hCG standard (Ovitrelle). Sample treatment types are listed under the graph (OFEL stands for open-faced endometrial layer, EP for hormonal stimulation and 5B stands for 5 blastoids); average hCG level ± standard error of mean is shown (for 2 independent experiments, each experiment using OFEL from 2 different donors). Statistical significance of grouped comparisons (ANOVA with Dunnett’s correction for multiple comparisons): ** P < 0.01, * P < 0.05, ns – not significant.

## Discussion

In this study, we used *in vitro* models that recapitulate key features of the human peri-and early post-implantation embryo development by growing human stem cell-derived blastoids with donor-derived endometrial epithelial cells as OFEL and on laminin coated surfaces, respectively. EEOs were re-seeded as OFELs to recreate the apical surface of the endometrial epithelium [8], while blastoids, representing a blastocyst-like structure composed of the three founding embryonic lineages, served as the embryonic component of the model [6]. Co-cultures were set up by transferring five blastoids onto either hormonally stimulated or unstimulated OFELs, and attachment and endocrine responses were assessed after 2 days [3]. To extend these observations beyond the attachment phase, we further employed a laminin-521–based post-implantation model that enables analysis of blastoid developmental progression in a defined extracellular matrix environment after attachment. This configuration allowed us to evaluate not only the physical interaction between the blastoid and endometrial surface, but also the functional endocrine and transcriptomic responses accompanying early implantation-like events.

Human blastocysts produce hCG prior to implantation in assisted reproduction settings [40], and the blastoids used in our model exhibited similar behaviour. Moreover, disruption of hCG–LHCGR signalling abolished embryo–endometrium attachment and subsequent developmental progression, identifying embryonic hCG as a key regulator of implantation competence. Transcriptomic analysis revealed 41 genes differentially expressed in both hormonally stimulated and unstimulated co-cultures, including CGA and CGB3/5/8. Notably, *CGB7* emerged as the only β-subunit gene significantly upregulated in the hormonally stimulated OFEL with attached blastoids, suggesting context-dependent regulation by both endometrial priming and embryo–maternal interaction. Whereas CGB1/2 are pseudogenes, the coding genes can be further classified as type I (CGB7) or type II (CGB3/5/8) [16], differing by the sequence of three amino acids [41]. Whereas *CGB3*, *CGB5*, and *CGB8* share conserved upstream promoter features, *CGB7* exhibits a clearly divergent promoter configuration, which aligns with evidence that its transcription is inducible by the p53 tumour suppressor, in contrast to the other family members. Previous studies have shown that CGB7 is expressed at lower levels than CGB3/5/8 during pregnancy [42] but may have distinct regulatory roles, particularly within the endometrial and decidual compartments [43–45]. Functional hCG is formed through heterodimerization of the common α-subunit (*CGA*) with any single coding CGB gene, thus, selective induction of *CGB7* is sufficient to support hCG production in this system. Given its sequence divergence from *CGB3/5/8*, *CGB7* expression may reflect an early, locally acting hCG signal during peri-implantation embryo–endometrial interactions rather than a systemic endocrine effect.

Recent advances in *in vitro* endometrial modelling allowed to investigate the molecular and cellular changes upon implantation and post-implantation of the embryo [20,21]. Consistent with the initiation of STB differentiation, our transcriptomic analysis revealed differential expression of established STB-associated genes, including *ERVW-1*, the CGB cluster, *CGA*, and *TBX3*, as previously reported [20,21]. Notably, our endometrial model captures the first 2 days following embryo attachment an earlier time point than those examined in recently published systems, which extend from 3 days to up to 14 days post-attachment. This restricted window, together with the use of an epithelial-only endometrial compartment, provides a focused view of the earliest transcriptional events accompanying embryo attachment, preceding more advanced trophoblast maturation and invasion.

To further distinguish embryo–maternal signalling during attachment from intrinsic embryonic developmental programs, we complemented the blastoid–OFEL co-culture system with a simplified post-implantation model based on laminin-521. In this minimal extracellular matrix environment, blastoids underwent progressive morphogenetic changes from day 2 to day 7 post-attachment, including structural expansion and lineage organization reminiscent of early post-implantation development. The system supported the emergence of specialized trophoblast-like populations, including hCGβ-positive syncytiotrophoblast-like cells and HLA-G-positive extravillous trophoblast-like cells, while maintaining distinct embryonic lineage identities marked by GATA3-positive trophectoderm-like derivatives and GATA4-positive hypoblast-like cells. Although this laminin-based system lacks maternal cellular components, the observed developmental progression indicates that key aspects of early trophoblast differentiation can proceed in response to extracellular matrix cues once attachment competence is established. These findings suggest that the embryo–maternal signalling events captured in the OFEL co-culture model likely represent an upstream phase that enables subsequent ECM-driven trophoblast maturation and morphogenesis. Together, the two complementary systems provide a modular framework in which early attachment-associated signalling can be studied separately from the intrinsic developmental programs that shape early post-implantation embryonic architecture.

The hCG immunoassay confirmed that blastoids secreted hCGα even prior to attachment, with secretion levels increasing further upon interaction with the endometrial layer. This pattern mirrors the physiological behaviour of human blastocysts, which secrete free hCGα in large amounts [16] and produce hyperglycosylated hCG (hCG-h) as the dominant form during early implantation [46] to promote trophoblast invasion and cytotrophoblast proliferation. In contrast, no hCG was detected in OFEL-only samples, irrespective of the hormonal stimulation.

To evaluate whether the hCG secreted in the co-culture model was biologically active, we performed a cell-based LHCGR activation assay that quantifies intracellular cAMP accumulation as a downstream readout of receptor engagement. Although normal and hyperglycosylated hCG differ in receptor affinity and signalling dynamics [14,15], our functional assay did not discriminate between these molecular variants. Therefore, to approximate the relative amount of bioactive hormone, we compared the dose-response curves obtained from serial dilutions of each sample to the reference curve of Ovitrelle (recombinant hCG), which was included as a positive control in every experiment. By assuming that the EC_50_ value of each sample corresponded to that of Ovitrelle, we expressed the data as Ovitrelle-equivalent hCG concentrations, allowing for standardized comparisons between experimental groups.

The functional hCG activity detected in the LHCGR assay closely mirrored the protein-level results obtained by immunoassay. In most samples containing blastoids alone, no measurable increase in cAMP was observed, whereas co-culture with OFELs consistently elicited receptor activation, indicating that the presence of an OFELs enhanced either the amount or the stability of bioactive hCG. The trend toward higher cAMP induction in hormonally stimulated OFEL-blastoid co-cultures, though not statistically significant, further supports a scenario in which endometrial hormonal priming facilitates productive embryo-maternal signalling. Together, these observations confirm that hCG secreted in the attachment model is not merely present at the protein level but also biologically competent to activate its canonical receptor, underscoring the physiological relevance of the blastoid-endometrium interaction reproduced *in vitro*.

Several previously reported *in vitro* models of human embryo-endometrium interaction have relied on trophoblast spheroids [47] or trophoblast cell line spheroids [48,49]. Other studies have used naïve pluripotent stem cell-derived blastoids, or human or primate blastocysts, yet co-cultured these with immortalized primary endometrial stromal cells [50], or extracellular matrix models [22,51], thus omitting the epithelial component which is the critical first point of contact for the implanting embryo. This model advances the field by integrating human donor-derived endometrial epithelial compartment with hPSC-derived blastoids that faithfully recapitulate the three founding embryonic lineages (EPI, TE, and PrE). While some of the previous systems employed similar cellular components [6] or even more complex endometrial part (*e.g*., epithelial and stromal assembloids [4,52]), our study additionally distinguishes itself through its focus on peri- and early post-implantation events, specifically transcriptomic profiling and hCG secretion as a sentinel biomarker of implantation success, offering a robust and quantifiable functional endpoint for future studies.

Because hCG secretion reflects both embryonic viability and cross-talk with the receptive endometrium, it represents a quantifiable informative readout to assess the effects of pharmacological or environmental agents on implantation dynamics. Furthermore, hCG, and especially its hyperglycosylated form, which was the dominant form of hCG in our model, has also been functionally implicated in implantation process [46,53]. The inclusion of a functional LHCGR assay further extends this paradigm, as it enables discrimination between mere hormone presence and biologically active signalling capacity, a critical distinction for understanding how chemical exposures or therapeutic agents might modulate early pregnancy processes. In this regard, our work bridges the gap between simpler attachment assays and more complex *in vivo* physiology, offering both mechanistic insight and experimental control.

Nonetheless, several limitations remain. The current endometrial component OFEL, while hormonally responsive and structurally accessible, represents a 2D simplification of the native endometrium. *In vivo*, the endometrium is a complex 3D tissue comprising multiple epithelial subtypes, stromal fibroblasts, vascular and immune components, and dynamic extracellular matrix interactions. To add to this, in the post⍰implantation model, blastoid differentiation occurs entirely in the absence of maternal support and without decidual assistance, taking place on a laminin⍰coated surface that further deviates from natural conditions. Moreover, extending the co-culture duration and spatial complexity could enable studies of decidualization, invasion depth, and maternal immune tolerance, capturing later implantation events currently beyond the scope of this platform.

Our findings demonstrate that embryo attachment initiates coordinated developmental and endocrine programs, and provide direct evidence that hCG secretion is dynamically regulated within the embryo–endometrial microenvironment. Importantly, hCG produced in these systems is not only detectable but biologically active, supporting a broader role for embryonic endocrine signalling. This study provides functional validation of embryo–endometrium crosstalk. This work defines a tractable framework to investigate early human implantation biology and offers a basis for studying mechanisms of implantation failure and the impact of external perturbations on early pregnancy.

## Ethics statement

EEOs were generated using endometrial biopsies obtained from healthy female donors in South Estonian Hospital (Võru, Estonia) under ethical permissions No. 330/M-8 (issued on 16.10.2020) and No. 364/M-9 (issued on 16.05.2022) by the Research Ethics Committee of the University of Tartu, Estonia. The use of human embryonic stem cells for generation of blastoids and their use in co-cultures was approved by ethical permits EPN #454/02 and #2011/745/31-3 granted to Prof. Fredrik Lanner and Prof. Ganesh Acharya at Karolinska Institutet, Sweden.

## Data availability

The mRNA sequencing raw data are available are available from the corresponding author upon request.

## Author roles

Conceptualization: DL, JJA, PD, AS, AS-L, AA, GA, FL. Methodology: DL, AS-L, AA, ADSP, IR, KK, HK, SR, MB, KH. Investigation: DL, AS-L, AA, ADSP, PIM, AR, IR, SR, MS. Writing (original draft): DL, AS-L, AA, PIM, HK. Writing (review and editing): all authors. Validation: DL, AS-L, AA, ADSP, PIM, AR, IR, SR. Formal Analysis: DL, AA, PIM, HK. Visualization: DL, AS-L, AA. Resources: DL, HK, FL, GA, PD, AS, AS-L. Data Curation: DL, AA Supervision: DL, AS, AS-L, AA. Project Administration: PD, AS, AS-L, FL, GA. Funding Acquisition: DL, PD, AS, HK, AS-L, FL, GA.

## Acknowledgements

We thank the patients who participated in this study for donating their tissues. We thank Annikki Löfhjelm for technical help with immunoassays, and Prof. Ago Rinken group members for the maintenance of the microplate reader platform. This study was partially performed at the Live Cell Imaging Core facility/Nikon Centre of Excellence, at Karolinska Institutet, Sweden, supported by the KI infrastructure council.

## Financing

This work was supported by the Estonian Research Council grants (PSG1082 and PRG1076); Centre for Innovative Medicine (CIMED), Region Stockholm no. FoUI-963306; Swedish Research Council grant no. 2024-02530 and Novo Nordisk Foundation grant no. NNF24OC0092384. DL was supported by the internal financing from the Institute of Chemistry, University of Tartu, Estonia (PLTKTARENG21). A.S. was supported by Horizon Europe (NESTOR, grant no. 101120075) and the Estonian Ministry of Education and Research Centres of Excellence grant TK214 name of CoE. HK was supported by the Sigrid Jusélius Foundation. MB was supported by Wenner-Gren foundation (grant no. UPD2022-0108). KH was supported by Estonian Research Council (PUTJD1253). PD and IR received financial support from the European Union (ERC, SAFER, 101124440). Views and opinions expressed are those of the authors only and do not necessarily reflect those of the European Union. Neither the European Union nor the granting authority can be held responsible for them.

## Supplementary Legends

***Supplementary Figure S1***. Gene ontology (GO) analysis of differentially expressed genes (DEGs) in hormonally and non-hormonally stimulated attachment models. **A**, GO analysis of biological processes based on 123 DEGs altered exclusively in the hormonally stimulated attachment model. **B**, GO analysis of biological processes based on 44 DEGs altered exclusively in the unstimulated attachment model.

***Supplementary Figure S2***. Pilot experiments in LHCGR activation assay: autofluorescence correction and confirmation of absence of 8-Br-cAMP interference. **A-C**, Kinetic curves showing changes in cAMP-sensor signal (ΔFRET) after addition of dilutions of Ovitrelle (**A**), Ovitrelle spiked into the hormone-containing (EP) open-faced endometrial layer (OFEL) medium (**B**), or EP-containing OFEL medium alone (**C**) onto the LHCGR-expressing cells. Note that high autofluorescence of the medium in the 530 nm optical channel is evident as decrease in ΔFRET. **D**, dose-response curves for Ovitrelle diluted in the assay buffer (full symbols) and for Ovitrelle spiked to the EP-containing OFEL medium after the subtraction of the medium-alone signal (empty symbols). **E**, comparison of negative logarithm of EC_50_ (pEC_50_) and top plateau values for the dose-response curves obtained in **D**. No statistically significant differences between the curve parameters were observed according to the unpaired t-test with Welch’s correction.

***Supplementary Table S1***. Differential expression analysis of transcriptomic data. **A**, 123 differentially expressed genes (DEGs) obtained for comparison between hormonally stimulated OFEL with blastoids *versus* hormonally stimulated open-faced endometrial layer (OFEL) alone. **B**, 44 DEGs obtained for comparison between non-stimulated OFEL with blastoids *versus* non-stimulated OFEL alone. In both sheets, the DEGs are ranked by binary logarithm of fold change (log (FC)).

***Supplementary Table S2***. Absolute (left) and normalized (right) concentration values for the secreted proteins measured in conditioned culture media using selective immunoassays. **A**, free hCGβ (absolute concentration units: pmol/L); **B**, free hCGα (absolute concentration units: pmol/L); **C**, hyperglycosylated hCG (absolute concentration units: pmol/L); **D**, total heterodimeric hCG (absolute concentration units: pmol/L); **E**, glycodelin (absolute concentration units: ng/mL).

