## Supplementary figures and images for "Functional profiling of human chorionic gonadotrophin in embryo peri- and post-implantation *in vitro* models"

### Supplementary Figure S1

#### Slide 1
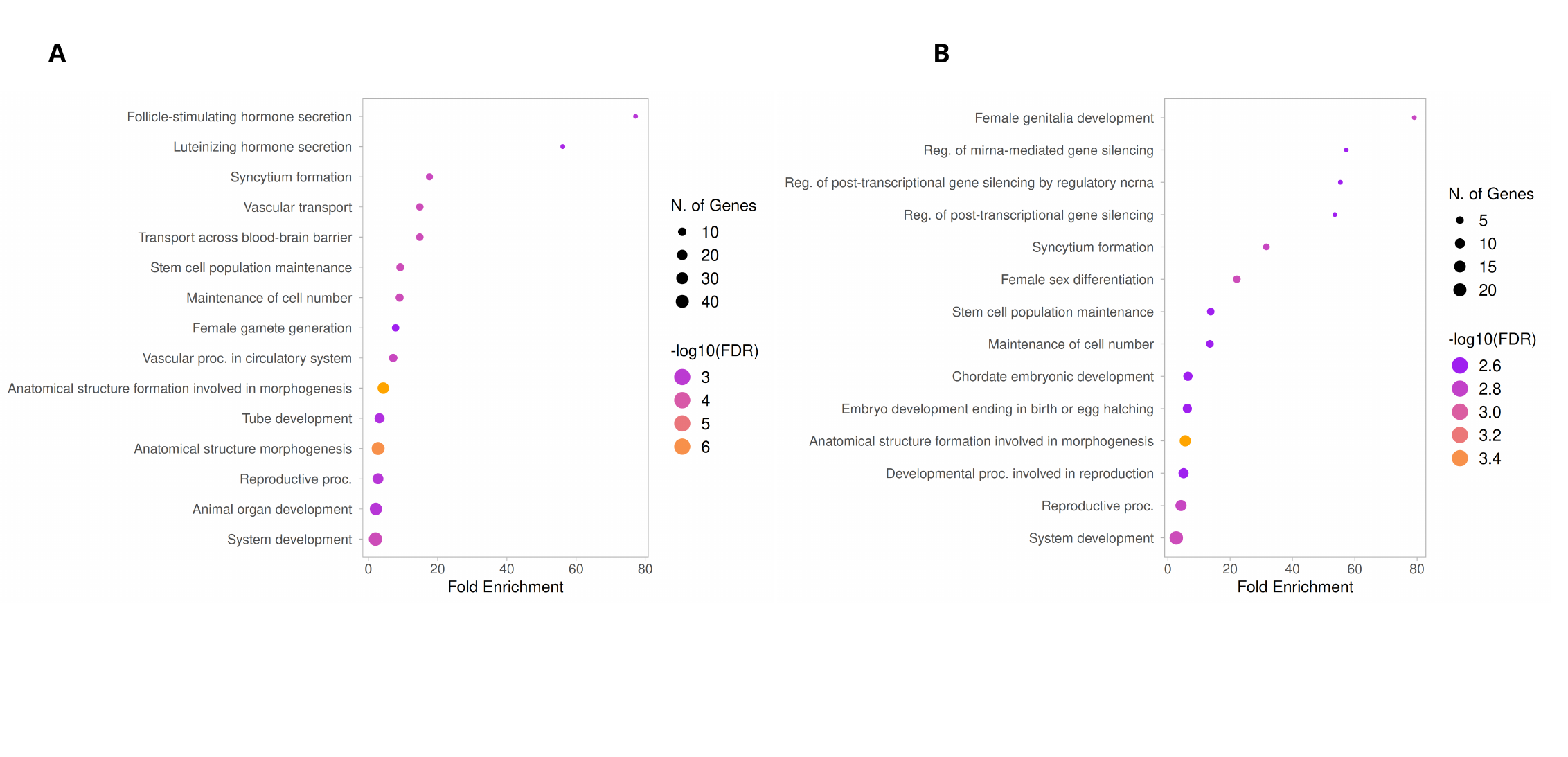

A
B

### Supplementary Figure S2

#### Slide 1
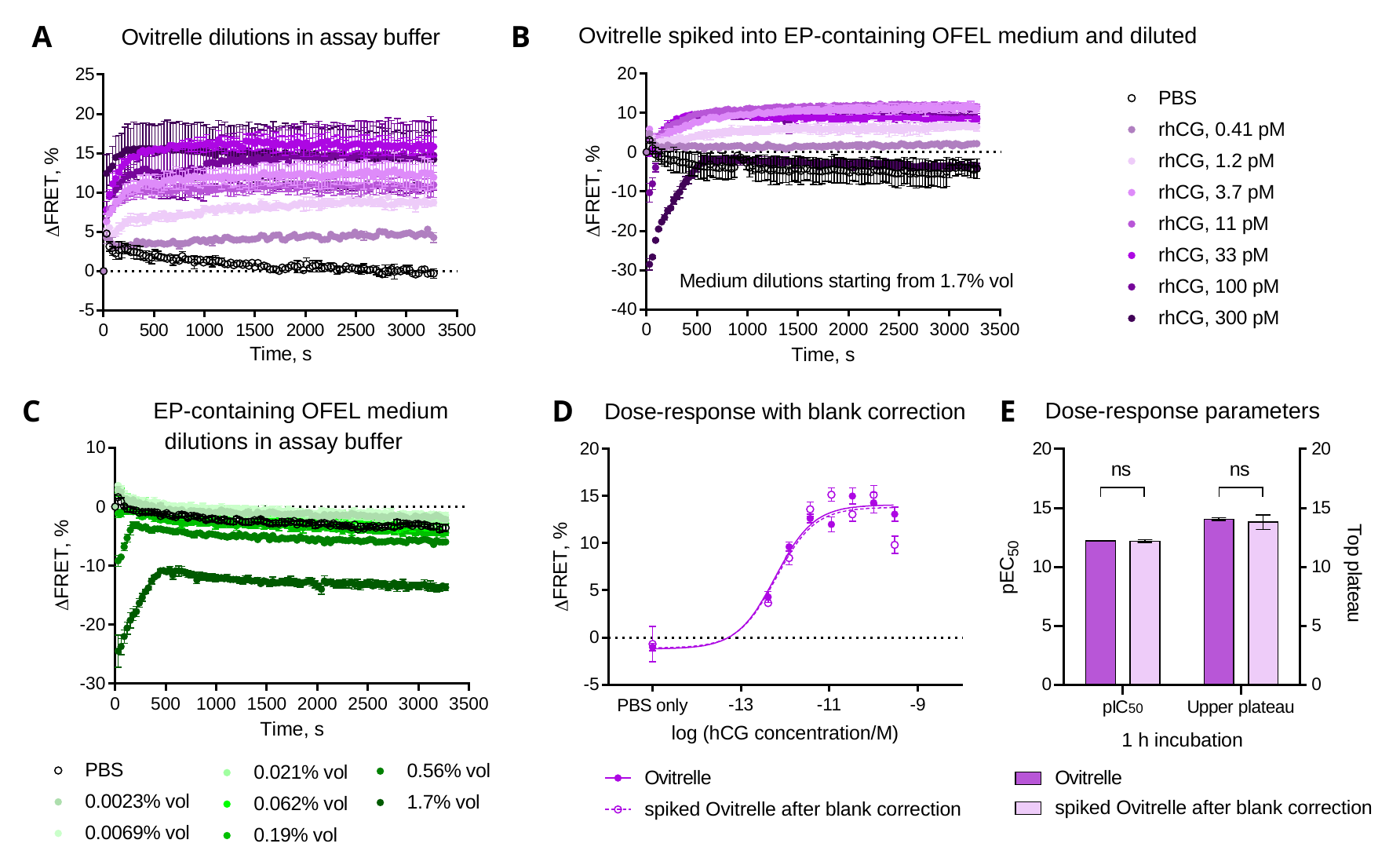

A
B
C
D
E
